# COVID-19 vaccination did not improve employee mental health: A prospective study in an early phase of vaccination in Japan

**DOI:** 10.1101/2021.09.02.21262808

**Authors:** Natsu Sasaki, Reiko Kuroda, Kanami Tsuno, Kotaro Imamura, Norito Kawakami

## Abstract

The effect of the COVID-19 vaccination as an individual-based preventive measure on mental health is largely unclear in the literature. The authors report a preliminary finding on whether vaccination effectively improves mental health among employees in Japan based on a prospective study (E-COCO-J). Of the total sample (N=948), 105 (11.1%) were vaccinated at least once at the follow-up survey (June 2021). There was no significant effect of vaccination on the change of psychological distress at baseline (February 2021) and follow-up (June 2021), after adjusting for gender, age, marital status, education, chronic disease, company size, industry, and occupation (healthcare workers or non-HCWs). Providing continuous mental health care for employees is important in an early vaccination phase.

## Manuscript

Deteriorated mental health is a major public health concern during the COVID-19 pandemic among working populations, including healthcare workers (HCWs) and non-HCWs, and the community overall ^1^. Vaccination is considered an effective individual-based preventive measure to control the COVID-19 epidemic ^2^. People adopted individual-based preventive measures, such as hand-washing, wearing masks, and others, to cope with fear and worry about COVID-19 ^3^. Using a COVID-19 contact tracking app was associated with better mental health ^4^. However, the effect of the COVID-19 vaccination as an individual-based preventive measure on mental health is largely unclear in the literature. We thus report a preliminary finding on whether vaccination effectively improves mental health among employees in Japan based on a prospective study.

The data were retrieved from the Employee Cohort Study conducted during the Covid-19 pandemic in Japan (E-COCO-J) ^5,6^. The Research Ethics Committee of The University of Tokyo approved this study (No. 10856-(2)(3)(4)(5)). We measured psychological distress using an 18-item scale of the Brief Job Stress Questionnaire (BJSQ) [possible range: 18 - 72] ^7^ at T1 (4 - 10 February 2021) and T2 (22 – 29 June 2021). We limited the analytic sample to individuals employed at both T1 and T2 without missing covariates. In Japan, priority vaccination for HCWs began on February 17, followed by people over 65 and people with chronic diseases. Vaccination programs started on June 21 in a few workplaces.

Of the total sample (N=948), 105 (11.1%) were vaccinated at least once at T2. Participants vaccinated at least once were more likely to be females (p<0.001), employed in a company with 300-999 employees (p=0.007), work in the medical and welfare industry (p<0.001), and attain college/vocational education (p<0.001). Proportions of vaccinated participants were higher for healthcare workers (HCWs) (n=56/103: 54.4%) than for non-HCWs (n=47/817: 5.8%) and participants with unknown occupation (n=2/28: 7.1%) (df=2, p<0.001). Differences between vaccinated and non-vaccinated participants in age, marital status, and chronic physical condition were non-significant.

The crude mean scores (standard deviations, SDs) of psychological distress at T1 and T2 were 41.8 (10.9) and 42.0 (11.9) for vaccinated participants and 41.2 (11.4) and 41.2 (11.6) for non-vaccinated participants, respectively, with no significant effect of having been vaccinated (Cohen’s d= 0.02, p=0.833 for the time x group interaction by repeated ANOVA). After adjusting for gender, age, marital status, education, chronic disease, company size, industry, and occupation (HCWs or non-HCWs), the estimated mean scores (standard errors, SEs) of psychological distress at T1 and T2 were 41.4 (1.7) and 42.4 (1.8) for vaccinated participants and 43.3 (1.2) and 43.4 (1.2) for non-vaccinated participants, respectively, with no significant effect (p=0.446).

Our study did not show improved psychological distress among participants vaccinated at least once compared to non-vaccinated individuals immediately after vaccination started in Japan. The findings suggest that vaccination for COVID-19 may not improve workers’ mental health, especially not worsened and sustained psychological distress during COVID-19 outbreaks ^5^. An effort to continuously monitor and improve workers’ mental health during the COVID-19 should continue despite vaccination.

A previous study reported that people who had higher stress are more likely to use individual-based preventive behaviors (e.g., washing hands) ^8^. Therefore, vaccinated participants would be expected to have higher stress. However, since we adjusted for psychological distress before the vaccination in our study, this expectation does not apply to our findings. Another possible reason is that the study was conducted in an early phase of vaccination in this country. People may not be fully convinced that the vaccination effectively reduces the risk of COVID-19 infection. In addition, employees’ mental health may be affected not only by a perceived risk of COVID-19 infection but also by social and economic problems due to the epidemic.

Several limitations should be addressed. The sample was small, comprising full-time employees recruited via an online platform in Japan. HCWs included employees working in non-clinical settings. Only a few participants were vaccinated, and some were vaccinated only once. Analyzing samples of fully vaccinated individuals in other countries and another phase of a pandemic may lead to different conclusions. In conclusion, providing mental health care for employees is important in an early vaccination phase.

## Data Availability

Data availability statement: The data that support the findings of this study are available from the corresponding author, NK, upon reasonable request.

## DISCLOSURE

### Approval of the research protocol

This study was approved by the Research Ethics Committee of the Graduate School of Medicine/Faculty of Medicine, The University of Tokyo, No. 10856-(2)(3)(4)(5).

### Informed consent

Online informed consent was obtained from all participants with full disclosure and explanation of the purpose and procedures of this study. We explained that their participation was voluntary, and they can withdraw consent for any reason, simply by not completing the questionnaire.

### Registry and registration number of the study/trial

N/A.

### Animal studies

N/A.

## Author contribution

NK was in charge of this study, supervising the process and of providing his expert opinion. NS and NK organized the study design and analyzed the data. Collaborators RK, KT and KI ensured that questions related to the accuracy or integrity of any part of the work were appropriately investigated and resolved. All authors participated in conducting the survey. NS and NK wrote the first draft of the manuscript, and all other authors critically revised it. All authors approved the final version of the manuscript.

## References

1. Cenat JM, Blais-Rochette C, Kokou-Kpolou CK, et al. Prevalence of symptoms of depression, anxiety, insomnia, posttraumatic stress disorder, and psychological distress among populations affected by the COVID-19 pandemic: A systematic review and meta-analysis. Psychiatry research. 2021;295:113599.

2. Thompson MG, Burgess JL, Naleway AL, et al. Interim estimates of vaccine effectiveness of BNT162b2 and mRNA-1273 COVID-19 vaccines in preventing SARS-CoV-2 infection among health care personnel, first responders, and other essential and frontline workers—eight US locations, December 2020–March 2021. Morbidity and Mortality Weekly Report. 2021;70(13):495.

3. Minisry of Health L, and Welfare. The results of the national mental health survey in the COVID-19 pandemic (Japanese). 2020; https://www.mhlw.go.jp/stf/newpage_15790.html. Accessed 12, July, 2021.

4. Kawakami N, Sasaki N, Kuroda R, Tsuno K, Imamura K. The Effects of Downloading a Government-Issued COVID-19 Contact Tracing App on Psychological Distress During the Pandemic Among Employed Adults: Prospective Study. JMIR Ment Health. 2021;8(1):e23699.

5. Sasaki N, Asaoka H, Kuroda R, Tsuno K, Imamura K, Kawakami N. Sustained poor mental health among healthcare workers in COVID-19 pandemic: A longitudinal analysis of the four-wave panel survey over 8 months in Japan. Journal of occupational health. 2021;63(1):e12227.

6. Sasaki N, Kuroda R, Tsuno K, Kawakami N. The deterioration of mental health among healthcare workers during the COVID-19 outbreak: A population-based cohort study of workers in Japan. Scandinavian journal of work, environment & health. 2020;46(6):639–644.

7. Inoue A, Kawakami N, Shimomitsu T, et al. Development of a short version of the new brief job stress questionnaire. Industrial health. 2014;52(6):535–540.

8. Newby JM, O’Moore K, Tang S, Christensen H, Faasse K. Acute mental health responses during the COVID-19 pandemic in Australia. PloS one. 2020;15(7):e0236562.

